# Effect of acupuncture on anxious symptoms in healthcare students with generalized anxiety disorder: A protocol for a randomized controlled clinical trial

**DOI:** 10.1101/2024.12.23.24319558

**Authors:** Cláudio Calixto Carlos da Silva, Marcos André de Matos, Sandro Rogério Rodrigues Batista

## Abstract

**Background:** The present study aimed to investigate the effect of acupuncture on generalized anxiety disorder (GAD) in healthcare students. On the basis of increasing prevalence of anxiety disorders in this population and the research gap regarding the use of acupuncture as a therapeutic intervention, this study aims to fill this gap and provide solid evidence on the effectiveness and safety of this practice.

**Methods:** The methodology will include a single-center, single-blinded, randomized, placebo-controlled clinical trial. A total of 87 patients diagnosed with generalized anxiety disorder will be enlisted and randomized to either manual acupuncture or sham acupuncture. Participants in both arms will undergo 30 minute-sessions once weekly until 10 sessions are completed over 4 months. The primary outcome measure will be assessed via 3 scales validated for Brazil: Generalized Anxiety Disorder Scale (GAD-7), The Beck Anxiety Inventory (BECK-A), The Penn State Worry Questionnaire (PSWQ) at baseline, 5 weeks and 10 weeks. All adverse events will be meticulously recorded and categorized by their time of onset and resolution, with appropriate clinical assessments provided for each patient.

**Discussion:** Acupuncture is expected to significantly reduce anxiety symptoms in participants. The results of this study will contribute to the evidence on the effectiveness and safety of acupuncture in treating anxiety in healthcare students, providing data that may influence clinical practices and future treatment guidelines.

**Trial Registration:** The Brazilian Registry of Clinical Trials: <u>RBR-9stskc6</u>. Registered 4 December 2024

## Introduction

The American Psychiatric Association defines generalized anxiety disorder (GAD) as excessive anxiety and worry about various events or activities lasting for more than six months. The intensity, duration, or frequency of worry is disproportionate to the actual likelihood or impact of the anticipated event, significantly impairing psychosocial functioning^1^. The World Health Organization (WHO) estimates that 264 million adults worldwide suffer from anxiety disorders, with Brazil having the highest prevalence at 9.3%^2^. Anxiety disorders are the most common mental health condition, affecting both individual well-being and public health^3,4^. Studies have shown that medical and healthcare students experience higher levels of anxiety, depression, and psychological distress than their peers do^5,6^. In Brazil, a meta-analysis by Pacheco^7^ revealed that trait anxiety was the most prevalent mental health issue among these students, affecting 89.6% of them. A broader meta-analysis revealed a 37.75% prevalence of anxiety among undergraduate students in Brazil, highlighting the high demands, competition, and stress inherent in academic life^8^.

The World Federation of Societies of Biological Psychiatry (WFSBP) recommends that treatment for anxiety disorders be individualized, considering factors such as illness severity, comorbidities, and previous treatments^9^. Therapeutic options include psychoactive drugs, psychotherapy, exercise, and other interventions. As first-line drug treatments for GAD, selective serotonin reuptake inhibitors (paroxetine, escitalopram, and sertraline) and serotonin and norepinephrine reuptake inhibitors (duloxetine and venlafaxine) are recommended.

However, patients with anxiety disorders have difficulty achieving full results when pharmacotherapy alone is used. Acupuncture, an integrative practice from traditional Chinese medicine, is widely used to treat mental health disorders. It has shown good tolerance and few side effects^10^. Acupuncture involves the insertion of fine needles into specific points on the body along meridians to promote health and prevent illness^11^.

A meta-analysis by Yang^12^ evaluated the effectiveness of acupuncture for treating GAD and demonstrated that acupuncture was more effective than control treatments, with good safety and tolerance. However, these studies were conducted in China, and full access to them through Brazilian databases is limited^13^. Dias^14^ conducted a randomized, placebo-controlled study in Brazil to assess the effects of electroacupuncture on stress symptoms in medical students. The study revealed no significant improvements in anxiety scales scores and did not use sham acupuncture groups.

An appropriate trial protocol is needed to investigate the effectiveness and safety of acupuncture in healthcare students in Brazil. Thus, we designed a single-center randomized controlled trial to observe and test the clinical effects of acupuncture for GAD, which is a study that has never been conducted before.

## Materials and methods

### Trial strategy

This is a randomized, placebo-controlled, single-blind, single-center clinical trial with healthcare students with a medical diagnosis of generalized anxiety disorder. The schedule for enrollment, interventions, and assessments is outlined in Fig 1, and the flowchart of the trial is shown in Fig 2.

**Fig 1.**
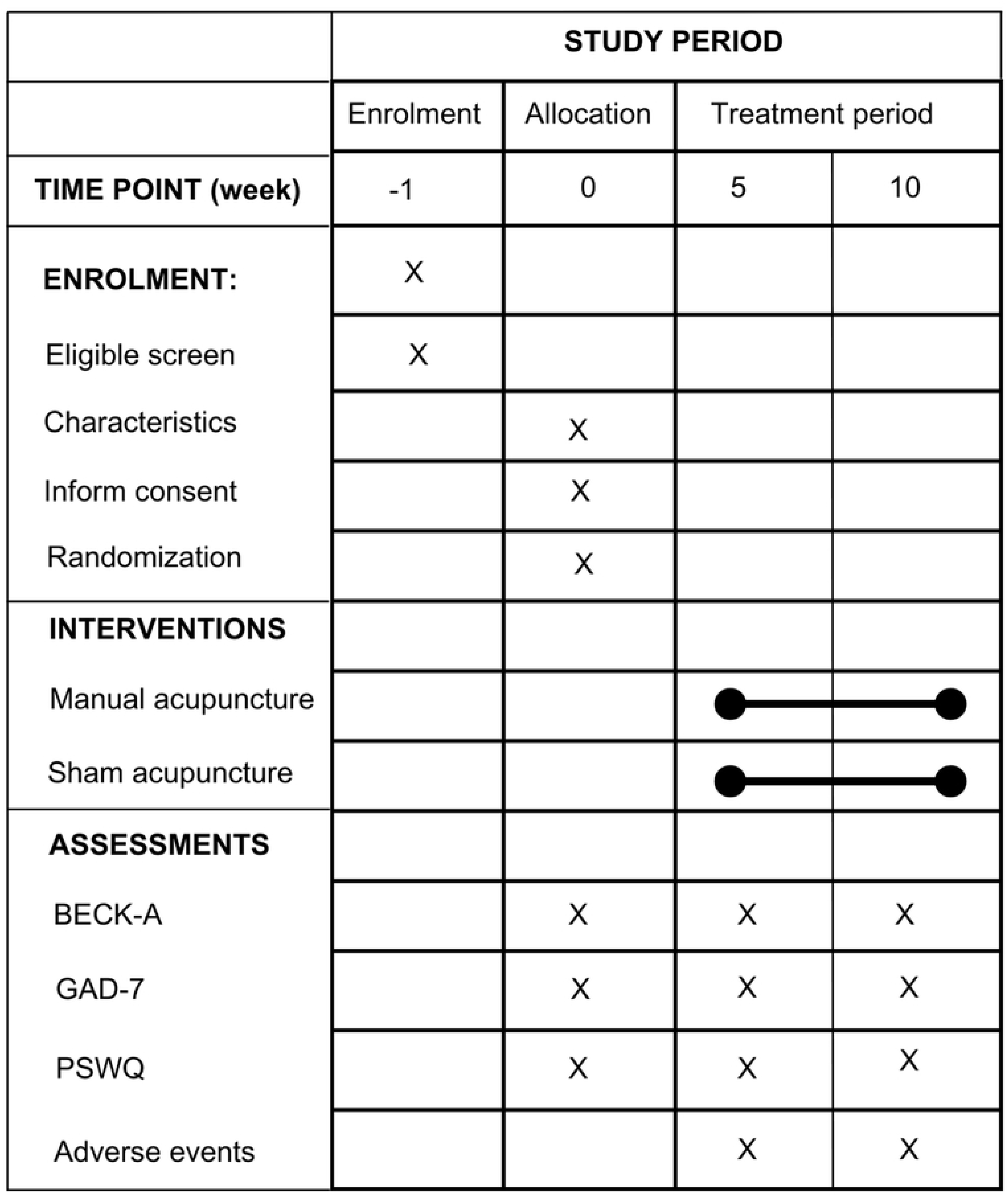
Trial SPIRIT schedule of enrollment, interventions, and assessment.

**Fig 2.**
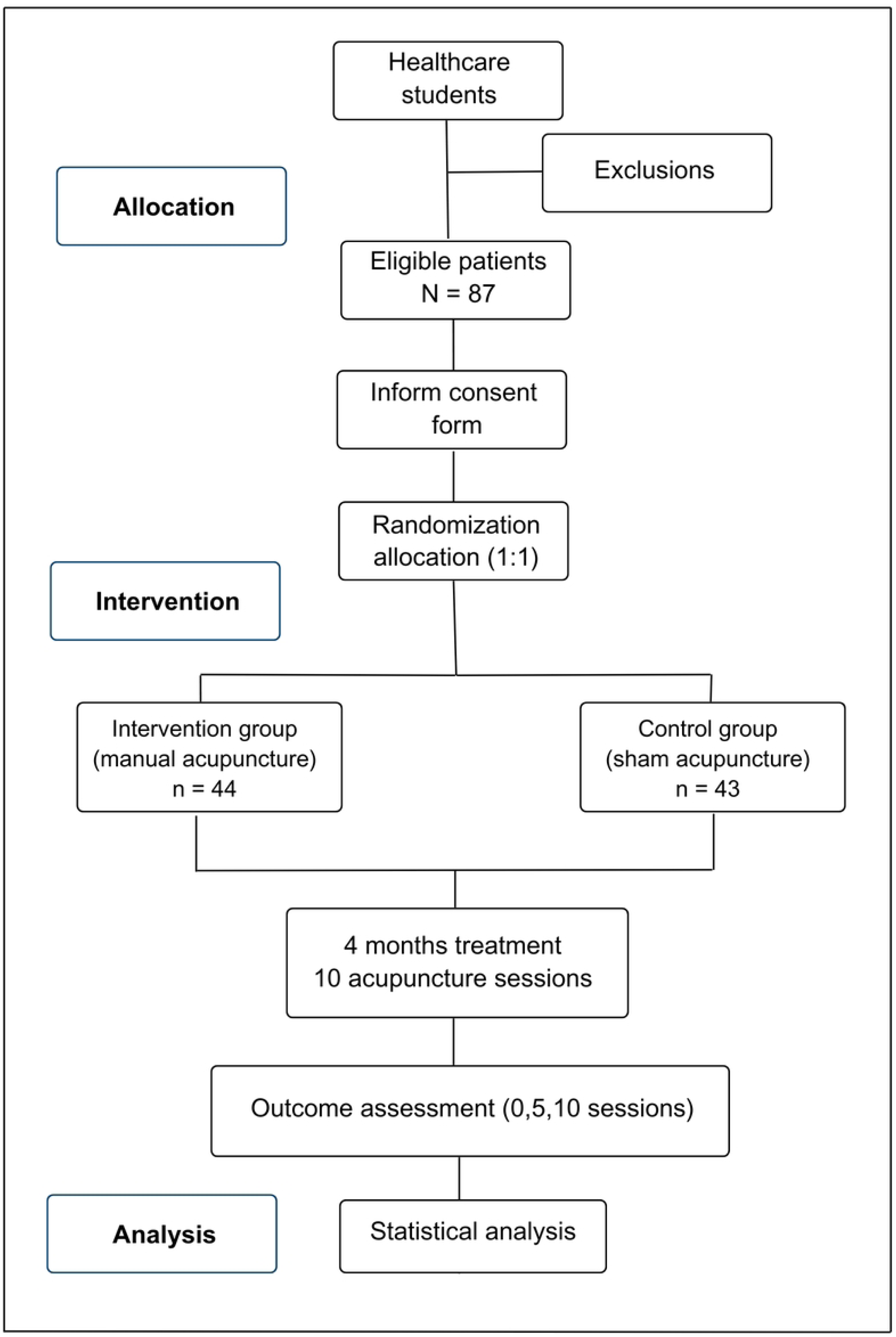
Trial Flowchart.

The study will follow the Consolidated Standards for Reporting Trials^15^ and the Guideline for Reporting Interventions in Acupuncture Clinical Trials^16^, CONSORT and STRICTA respectively. The checklist outlining these standards is provided in the S1 and S2 Appendix.

The randomized controlled clinical trial (RCT) will be conducted at an Integrative and Complementary Practices outpatient clinic of a health institution in Goiânia, Goiás between March 2025 and June 2025.

### Patients

#### Study population

This trial is slated to commence in March 2026, concluding in June 2026, with a target enrollment of 87 healthcare students with generalized anxiety disorders. To ensure the precision of the results of this trial, patients who meet the following eligibility criteria will be included. Eligible subjects shall be apprised of the prospective benefits and potential risks of participation, with written Participation and Consent Form (S3 Appendix) obtained to guarantee voluntary enrollment. Wide publicity will be carried out through the Teams platform and via WhatsApp for all healthcare students at the educational institution. Once the inclusion and exclusion criteria have been met, students will be contacted via telephone/WhatsApp and invited to participate in the study. Those who agree to participate will have the initial assessment scheduled. As soon as the student, previously scheduled by phone/WhatsApp, attends the health institution, will be sent to the outpatient clinic.

#### Diagnostic criteria

The diagnosis of generalized anxiety disorder followed the The Diagnostic and Statistical Manual of Mental Disorders, Fifth Edition^1^.

#### Inclusion criteria

1. Meet diagnostic criteria according to the Fifth Diagnostic and Statistical Manual of Mental Disorders (DSM-V) criteria for GAD;
2. Standard treatment for GAD (psychotherapy and/or first-line psychiatric medications: paroxetine, sertraline, escitalopram, venlafaxine or duloxetine);
3. Symptoms of generalized anxiety disorder (generalized anxiety disorder scale - GAD-7) > 10;
4. Healthcare students regularly enrolled in the educational institution, over 18 years of age.

#### Exclusion criteria

1. Current psychosis or psychosis within the last 6 months;
2. Substance dependence within the last 6 months;
3. Severe depression that is significantly more relevant than GAD;
4. Pregnancy or postpartum women;
5. Treatment with acupuncture within the last 30 days;
6. Intolerance to acupuncture (needle phobia)
7. Healthcare students currently enrolled in the discipline of acupuncture and medical homeopathy;
8. Healthcare students with less than 1 year since the start of their degree program.

#### Criteria for withdrawal and termination

In accordance with ethical principles that protect patient autonomy, participants have the right to withdraw from the study at any time and for any reason. If a participant chooses to withdraw, the reason must be carefully documented and considered as a factor in the decision to terminate the trial. All primary data related to study withdrawals and terminations will be kept confidential until the completion of the trial.

#### Randomization procedure and allocation concealment

A total of 87 healthcare students will be randomly allocated through complete randomization in a 1:1 ratio via the website (https://www.random.org/) to the treatment arm (manual acupuncture group) or the control arm (sham acupuncture group).

#### Blinding

The group assignments will be kept concealed from the patients. The control group will undergo sham acupuncture with a non-penetrating device designed to resemble real acupuncture needles, ensuring the blinding (see Fig 3). Due to the inherent nature of acupuncture, it is not possible to blind the practitioners.

**Fig 3.**
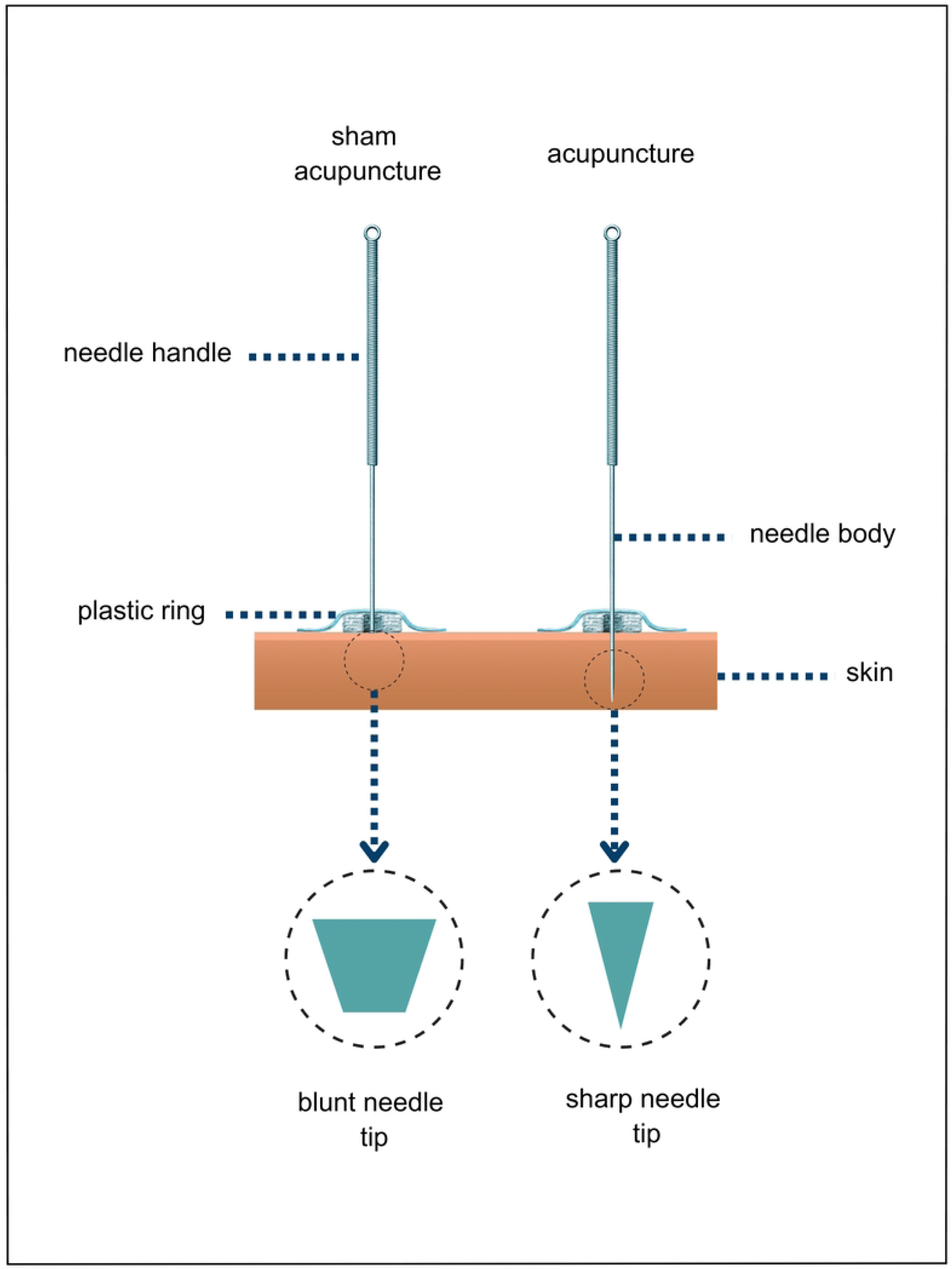
Scheme of acupuncture and sham acupuncture needles.

#### Interventions

Healthcare students will undergo 30 minute-sessions once weekly until 10 sessions are completed over 4 months. Acupuncture will be performed by an acupuncturist with more than 4 years of experience.

#### Treatment group

The needles will be inserted at an angle between 30° and 60°, with retention during the session without manipulation. The depth of insertion will be in accordance with the location of the point and the tension of the patient’s skin and subcutaneous tissue, generally 1 to 1.5 cm, with stimulation of rotation or pistoning until DeQi is obtained. DeQi will be considered as a feeling of pressure, heaviness, tingling, distension or pain around the needle insertion site.

In every session, each patient will receive a total of 10 needles. Patients will be instructed to lie in the supine position and will wear an individual blindfold and if they wish, they can fall asleep. Once the acupuncture needles are inserted, the patient will be left alone until the session ends.

Sterile, disposable stainless steel acupuncture needles measuring 30 mm by 0.25 mm (Dongbang, South Korea) will be used. The needles will be inserted into specific points according to clinical evidence on generalized anxiety disorder and the author’s knowledge of acupuncture. The choice of points will not take into account the individual syndromic diagnosis according to traditional Chinese medicine. The selected points were as follows: Yintang, HT-7 (Shenmen) bilateral, GV-20 (Baihui), PC-6 (Neiguan) bilateral, LR-3 (Taichong) bilateral, LI- 4 (Hegu) bilateral. The names and locations of the acupuncture points, shown in S1 table, are those described in accordance with the WHO international standard nomenclature of acupuncture^17^ and are based on the Acupuncture Atlas^18^.

#### Control group

The control group (sham acupuncture) will be treated with nonpenetrating needles by the same acupuncturist as the intervention group. When the tip of the needle touches the patient’s skin, it retracts into itself without piercing the patient’s skin.

Streitberger brand nonpenetrating needles will be used, with a total of 10 needles per patient allocated in the same location as the points used in the intervention group and with a 30-minute dwell time. In all the sessions, all the participants will wear an individual blindfold.

#### Primary outcomes

The primary outcome will be the statistically significant reduction in anxiety symptoms in the control group compared with the group undergoing sham acupuncture after 5 and 10 sessions.

Tree data collection instruments will be applied to those eligible before starting acupuncture sessions, namely:

1. The Beck Anxiety Inventory (BECK-A) is a self-report scale used to measure the intensity of anxiety symptoms. Twenty-one items, which are answered on a scale ranging from 0-3, have been validated for Brazil by Cunha^19^.
2. The Generalized Anxiety Disorder Scale (GAD-7) is a scale used to assess levels of anxious symptoms and is a four-point Likert-type scale (0-3 points), with seven items and scores ranging from 0-21 points, with 10 being the general cutoff point stipulated to attest to the significant presence of GAD symptoms. This scale has been validated for Brazil by Moreno^20^.
3. The Penn State Worry Questionnaire (PSWQ) is a scale used to assess excessive worry, with sixteen items answered on an agreement scale ranging from 1-5; this scale has been validated for Brazil by Castillo^21^.

After 5 sessions, all patients will undergo assessment with the 3 scales in the same way as the first assessment. After 10 sessions all patients will undergo assessment with the 3 scales in the same way as in previous assessments (Fig 2).

#### Secondary outcomes

The secondary outcomes will include the construction of scientific evidence on the use of acupuncture for generalized anxiety disorder and the formulation of recommendations on the basis of scientific evidence aimed at promoting mental health care students. The findings will be propagated through academic conferences and peer-reviewed publications.

#### Data gathering and administration

At screening, study personnel initiated collection of data on individuals meeting eligibility criteria, encompassing demographics, work, academic and clinical characteristics.

#### Patients’ safety and adverse events

The most common adverse reactions are vasovagal hypotension, painful sensations and hematoma at the needle insertion site. All patients will be monitored and noted in each acupuncture session, and a clinical assessment relevant to each patient will be provided. In the case of other reactions, students will be referred to other medical specialties at the health institution itself.

#### Sample size

To calculate the sample size, the population of healthcare students of the educational institution targeted by the study (N = 3667) was considered, with 36.1% of this population meeting the inclusion criteria. To this end, the classic procedure described by Lwanga and Lemeshow^22^ was used to estimate: n = N.Z².P (1 - P) / Z².P. (1 - P) + E².(N - 1)

“n” according to the equation where n = calculated sample; N = population; Z = standardized normal variable associated with the confidence interval; P = estimate of the proportion in the sample; and E = sampling error. Therefore, a 95% confidence interval, 5% sampling error and a 10% loss estimate were used. Therefore, the sample size for this study was 44 students in the intervention group and 43 students in the control group, totaling 87 students in the sample.

#### Statistical analysis

The data will be analyzed via the statistical package SPSS (Statistical Package for Social Science), version 26, with a significance level of 5% (p < 0.05). To characterize the sociodemographic, work, academic and clinical profiles of patients, descriptive statistics, such as the median, mean, standard deviation, minimum and maximum for continuous variables, and the absolute frequency and relative frequency for categorical variables, will be used. The distribution of the patients’ profiles in the intervention and control groups will be tested via Pearson’s chi-square test and Student’s t test.

The choice between parametric and nonparametric tests will be determined after applying the Shapiro-Wilk normality test. The comparison of anxiety symptoms measured by the BECK-A, GAD-7 and PSWQ before, during and after the intervention in the intervention and control groups will be carried out via analysis of variance (factorial ANOVA) for repeated measures.

#### Trial status

The trial will start recruiting participants in January 2026.

### Declarations

#### Ethics approval and consent to participate

The project received approval from the Research Ethics Committee (CEP) of the Federal University of Goiás (UFG) under protocol number 7.025.751.

All patients will receive the free and informed consent form (ICF) for reading, explaining and signing if they agree and agree to participate, with the guarantee of privacy and confidentiality of the information obtained. At any time, any patient may withdraw their consent to participate in the study without causing any harm to their care at the service. The ICF will comply with current Brazilian regulations in all ethical-legal aspects, that is, Resolution 266/12, which addresses studies with human beings.

Notably, all the material to be used will be disposable, and we will follow the recommendations for biosafety, patient safety, infection control in the health services and waste management and disposal.

## Discussion

Generalized anxiety disorder (GAD) is a prevalent mental health condition affecting healthcare students, who are particularly vulnerable due to academic pressure and professional expectations^5,6^. Insufficient scientific evidence exists regarding the efficacy of acupuncture for GAD in this population, especially in Western countries^12, 13^. This clinical trial is proposed as the first placebo-controlled study in Brazil focusing on healthcare students diagnosed with GAD and treated with manual acupuncture. Addressing this gap is crucial for developing non-pharmacological interventions that are both effective and accessible.

In generalized anxiety disorder (GAD), multiple neurotransmitter systems beyond serotonin appear to be implicated. These include norepinephrine, glutamate, and cholecystokinin, all of which contribute to the dysregulation of arousal and sleep, resulting in restless and unsatisfactory sleep^23^. Conventional pharmacological treatments, including anxiolytics and antidepressants, often pose risks such as dependency, cognitive impairment, and withdrawal symptoms^9^. In contrast, acupuncture has shown promise in modulating brain function, improving cerebral blood flow, and regulating neurotransmitters related to anxiety and stress responses. Studies suggest that acupuncture can reduce anxiety symptoms and promote autonomic balance, making it a safe, natural alternative to pharmacological therapies^10,12^.

This clinical trial aims to generate robust evidence supporting acupuncture as a viable intervention for GAD in healthcare students. The findings could inform mental health policies and provide a foundation for future research in this field. Results will be disseminated through presentations at healthcare and educational institutions, as well as publications in peer-reviewed journals, to foster knowledge exchange and strengthen mental health support for undergraduate students.

### Limitations

Despite the potential benefits of this clinical trial, several challenges and limitations must be acknowledged. One significant challenge is the recruitment and retention of participants. To ensure adequate enrollment, extensive outreach efforts through institutional communication platforms such as TEAMS and WhatsApp groups will be employed. Maintaining participant adherence to the study protocol is also crucial. To address this, regular communication and educational reinforcement on the importance of protocol compliance will be implemented to minimize dropout rates.

Ensuring methodological rigor through standardized interventions and effective blinding remains a priority. This trial will employ sham acupuncture as a placebo control, allowing participants to experience a similar tactile sensation^24^. This strategy aims to reduce bias and enhance the study’s validity. Additionally, anxiety levels will be measured using scales validated in Brazil to ensure the results are applicable to the local population.

However, this study has certain limitations. Notably, a follow-up assessment was not included, which restricts the ability to evaluate long-term outcomes. Furthermore, uncontrolled external factors, such as varying levels of academic stress and personal life changes among participants, may impact the results and introduce variability that is difficult to control.

## Data Availability

No datasets were generated or analysed during the current study. All relevant data from this study will be made available upon study completion.

## List of abbreviations

BECK-A: Beck Anxiety Inventory
CONSORT: Consolidated Standards for Reporting Trials
DSM-V: Fifth Diagnostic and Statistical Manual of Mental Disorders
GAD: Generalized Anxiety Disorder
GAD-7: Generalized Anxiety Disorder Scale
ICF: Informed Consent Form
PSWQ: Penn State Worry Questionnaire
RCT: Randomized Controlled Clinical Trial
STRICTA: Guidelines for Reporting Interventions in Acupuncture Clinical Trials

## Availability of data and materials

Following the publication of this study, the authors will communicate the trial results to participants, healthcare professionals, the public, and other relevant groups. The data are available upon reasonable request and with the permission of the authors.

## Author contributions

**Conceptualization:** Marcos Matos, Sandro Batista.

**Data curation:** Cláudio Silva.

**Formal analysis:** Cláudio Silva.

**Funding acquisition:** Cláudio Silva.

**Methodology:** Cláudio Silva.

**Project administration:** Cláudio Silva.

**Resources:** Cláudio Silva.

**Supervision:** Marcos Matos, Sandro Batista.

**Validation:** Cláudio Silva, Marcos Matos, Sandro Batista.

**Writing - original draft:** Cláudio Silva

**Writing - review & editing:** Cláudio Silva, Marcos Matos, Sandro Batista

## Acknowledgements

Not applicable.

## Supporting information

**S1 Appendix. CONSORT 2010 Checklist**

**S2 Appendix. SPIRIT 2013 Checklist**

**S3 Appendix. Informed Consent Form S1 File (PDF)**

**S1 Table. Allocation of acupoints and non-acupoints (PDF)**

